# Estimating COVID-19 infection fatality rate in Mumbai during 2020

**DOI:** 10.1101/2021.04.08.21255101

**Authors:** Murad Banaji

## Abstract

The aim of this piece is to provide estimates of the infection fatality rate (IFR) of COVID-19 in Mumbai during 2020, namely the fraction of SARS-CoV-2 infections which resulted in death. Estimates are presented for slums and nonslum areas, and for the city as a whole. These are based largely on the city’s official COVID-19 fatality data, seroprevalence data, and all-cause mortality data. Using recorded COVID-19 fatalities in the numerator, we obtain IFR estimates of 0.13%-0.17%. On the other hand, using excess deaths we obtain IFR estimates of 0.28%-0.40%. The estimates based on excess deaths are broadly consistent with the city’s age structure, and meta-analyses of COVID-19 age-stratified IFR. If excess deaths were largely from COVID-19, then only around half of COVID-19 deaths were officially recorded in the city. The analysis indicates that levels of excess mortality in excess deaths per 1000 population were similar in the city’s slums and nonslum areas. On the other hand the estimated ratio of excess deaths to official COVID-19 deaths in the slums was much higher than in nonslum areas, suggesting much weaker COVID-19 death reporting from the slums.

## 1. Introduction and main results

Arriving at estimates of the infection fatality rate associated with a COVID-19 epidemic presents challenges associated with estimating both the numerator (fatalities) and the denominator (infections). Given widespread evidence of COVID-19 death underreporting, both in India [1] and worldwide [2], taking recorded COVID-19 deaths at face value can introduce a major downward bias into IFR estimates.

Although Mumbai’s data is far from complete, the city has seen several seroprevalence surveys, and some all-cause mortality data is available. Death recording in the city is sufficiently stable to arrive at credible estimates of excess mortality during the pandemic.

The main results here are:

1. Estimates of COVID-19 IFR based on excess deaths during 2020, in the range 0.28%-0.40%, tally broadly with expectations from international data, for example from the meta-analyses in O’Driscoll *et al* [3] and Levin *et al* [4].
2. While COVID-19 IFR was probably somewhat lower in the slums (likely as a result of a younger population), this difference was not as stark as official fatality data suggests.
3. Undercounting of COVID-19 deaths was probably considerably higher in the slums than in nonslum areas. This likely explains the considerably higher ratio of excess deaths to official COVID-19 deaths during April-July 2020, when there was a major slum surge, as compared to the later part of 2020.

## 2. Basic notions

**Seroprevalence surveys** (“serosurveys” for short) to estimate the prevalence of antibodies to SARS-CoV-2 provide the main data used to estimate levels of prior infection. Mumbai has seen several serosurveys but the two carried out in July 2020 and August 2020 by the city corporation in collaboration with the Tata Institute of Fundamental Research [5, 6] were carefully planned and are accompanied by useful technical information. These are referred to as the city’s first and second serosurveys. The first one forms the cornerstone of the analysis here.

**Sensitivity and specificity** of antibody tests to prior infection is important to consider when interpreting serosurvey results. Given some number *k* of infections all occurring on a given day, we can ask how many of these would be picked up using some given test *n* days later. We expect sensitivity to peak two to three weeks after infection, and then decline; however the speed of the decline depends strongly on the test used [7]. In particular, the test used in Mumbai’s first and second serosurveys has fairly rapidly declining sensitivity.

**Prevalence** is used to refer to the fraction of the population who have been infected by SARS-CoV-2 and “recovered”. Since estimates of prevalence come from serosurveys, the time to recovery is defined as the time to seroconversion. The **infection rate** refers to the number of SARS-CoV-2 recoveries plus deaths as a fraction of the total population. Note that, given the possibility of reinfections, an infection rate of 50% does not necessarily imply that 50% of the population has been infected, and a value of over 100% is not in itself absurd. We assume that reinfections were relatively few by the time of the first serosurvey (July, 2020), so that prevalence is a good estimate of the infection rate.

**Infection fatality rate (IFR)** is the ratio of deaths to total infections, assuming that all those infected have either recovered or died. In practice, if we ignore deaths in the denominator this makes a very marginal difference to the estimates. For example, if there have been 3 deaths for every 1000 recoveries, ignoring the deaths in the denominator changes IFR from 0.299% to 0.3%.

**Naïve IFR** refers to any IFR estimate which ignores possible fatality underreporting and uses *recorded* fatalities in the numerator. Naïve IFR calculations are useful; but it is important to flag up the risk of downward bias if naïve IFR is used as a proxy for IFR.

**Fatality delay** refers to the delay used when estimating naïve IFR. If we have an estimate of prevalence on a given date and we assume a fatality delay of *n* days, then we would use recorded fatalities *n* days later when computing naïve IFR. The fatality delay needs to take into account possible delays in fatality recording. This is important, because Mumbai is known to have had major fatality recording delays and data reconciliations [9]. For example, suppose we estimate seroprevalence from an IgG antibody test, and suppose that IgG seroconversion typically occurs 14 days after symptom onset [8]; then using a fatality delay of 7 days is equivalent to assuming that deaths are typically *recorded* 21 days after symptom onset.

**Excess-deaths based IFR** or “**eIFR**” for short refers to the ratio of excess deaths to infections during a particular period.

**Undercount factor**. This refers to the ratio of excess-deaths based IFR and naïve IFR (in a given population during a given period). It can be regarded as the ratio of “true” pandemic mortality to “official” pandemic mortality. The terminology is not meant to imply that most or all excess deaths were necessarily COVID-19 deaths, although this may well be the case. Data from across India indicates huge undercount factors during the pandemic. In this context, Mumbai’s factor of around 2 is relatively low.

## 3. All-cause mortality in Mumbai during 2020

Yearly death registrations from 2015-2020, along with data on population, births and infant mortality, are available in a “Vital Statistics” report by the city corporation [10]. The key data from this report is in Table A.4 in Appendix A. During 2015-19 there was a marginally declining linear trend in yearly death registrations. 2019, however, saw 2.7% more death registrations than 2018. According to government data from 2014 onwards Maharashtra saw death registration coverage above 94% with complete coverage in 2018 and 2019 [11]. We assume that during 2017-19, Mumbai saw complete death registration.

Monthly death registration data for 2020, reported according to date of death, and partial monthly data for 2017-19 and 2021 is available at [12]. The data itself is given in Table A.5 in Appendix A. The availability of monthly registration data expands the scope of the analysis possible, raising the hope that we may be able to separately estimate fatality rates in the slums and nonslum areas, by aligning mortality data and seroprevalence data from the slums and nonslum areas.

To set mid-point expectations for monthly death registrations during 2020 we take averages over 2017-19. Relative to this baseline, monthly excess death registrations are plotted in Figure 1. Death registrations were around double expected during May 2020, about 70% higher than expected in June, and then plateaed at about 30% higher than expected from July to October.

**Figure 1:**
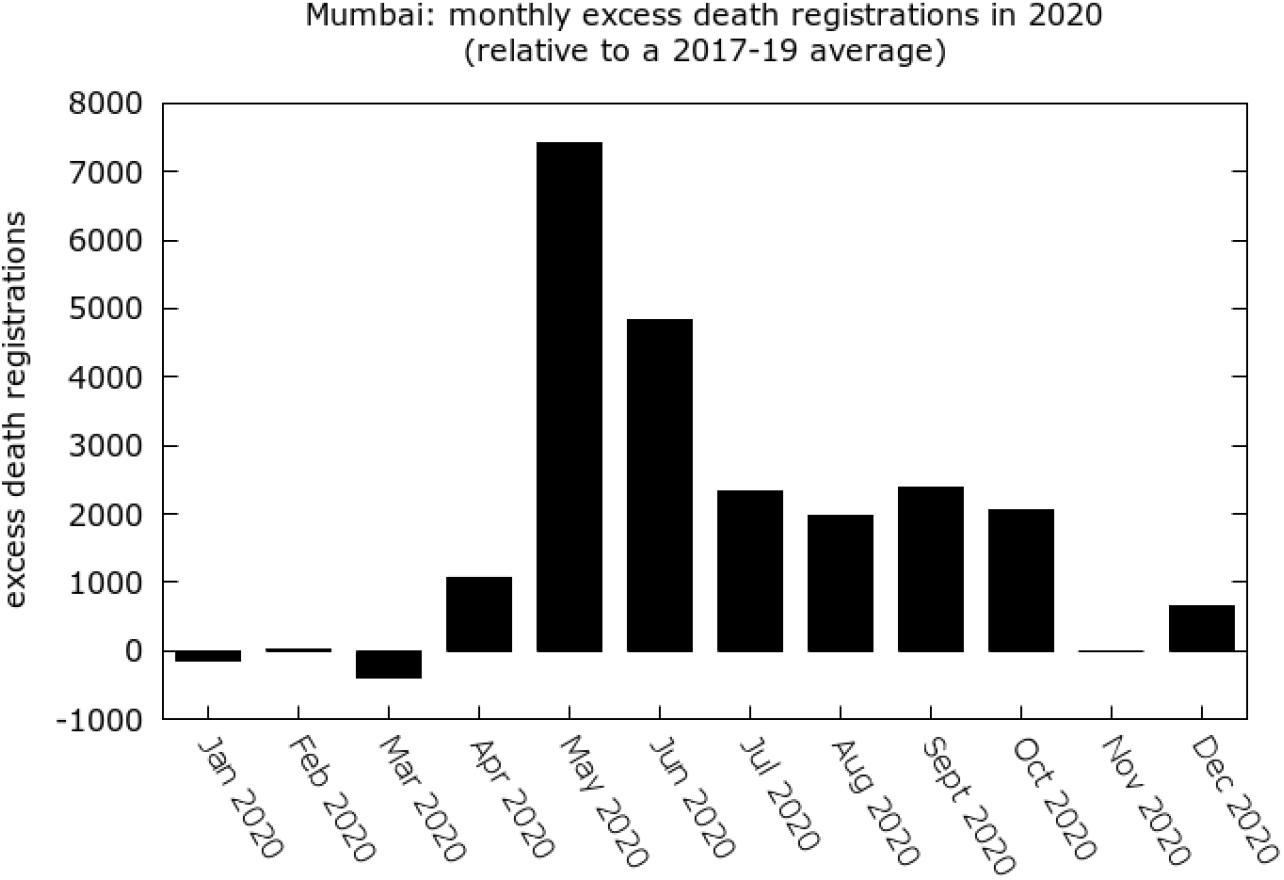
Monthly excess death registrations during 2020 in Mumbai relative to a 2017-19 average.

During April-December 2020, there were 22,753 excess death registrations, equivalent to a rise of 34% above expected values for these 9 months. On the other hand, Mumbai saw 11,116 recorded COVID-19 deaths in 2020. Thus, for each recorded COVID-19 death in 2020, the city saw close to two excess death registrations. If we ignore possible fluctuations in death registration during the pandemic, the undercount factor in the city was around 2 during 2020. This gross factor, however, ignores an important change: during April-July 2020, the undercount factor was around 2.5 (15,671 excess deaths and 6,344 recorded COVID-19 deaths), while during August-December 2020 it dropped to around 1.5 (7,082 excess deaths and 4,766 excess deaths). This fall coincided with the decline of the city’s huge slum-wave.

*Remark*. Although we do not consider 2021 data here, we remark that the undercount factor remained close to 2 during January-May 2021, although possible delays in official COVID-19 death recording and in death registration mean that this could shift in either direction.

There is evidence that death registration coverage fell and/or some forms of mortality fell during 2020. For example:

1. There was a drop in death registrations about 5% below expectations during March 2020, visible in Figure 1.
2. Registered births in 2020 fell by 23%, and infant deaths fell by 33%, relative to the previous five year average. These shifts are unlikely to reflect a shift in fertility, but could reflect a drop in population as a consequence of the migrant efflux from the city, or disruption to birth and death registration, or all of these.
3. According to the Hindustan Times [13], deaths on Mumbai’s suburban railway lines dropped by 1575 (58%) from 2,691 in 2019 to 1,116 in 2020 (no data is given prior to 2019). According to the Times of India [14], deaths on Mumbai’s roads dropped by 149 (36%) from 413 in 2019 to 264 in 2020 (no data is given prior to 2019).

In the light of this data, in later simulations we allow for some fluctuations in baseline mortality and a possible dip in death registration coverage. If, for example, there were *m* expected deaths during some period of 2020, *n* registered deaths during this period, and an assumed drop in registration coverage from 100% to (100 *− k*)%, we would then obtain an estimate of

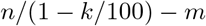

excess deaths during this period.

## 4. Estimating COVID-19 infections in Mumbai by the time of the first serosurvey

Mumbai’s first serosurvey [5] surveyed residents 12 years or older in three of the city’s 24 wards (F/N, M/W and R/N) for IgG antibodies to SARS-CoV-2. With an approximate mid-point of July 8, 2020, it returned seroprevalences of 54.1% (52.7% – 55.5%) in the slums surveyed, and 16.0% (14.8% – 17.2%) in the nonslum areas surveyed. These values were following reweighting to account for demographic characteristics (the raw values, inferred from Tables e2 and e6 in [5] were 57.0% and 15.7%), but were not corrected for test sensitivity or specificity of the assay used. The 95% CIs given by the authors reflect possible sampling errors.

According to 2011 data (given in Appendix A) 52.5% of the city lives in the slums. Thus slum and nonslum seroprevalence values of 54% and 16% respectively, if they held citywide, would imply 36% seroprevalence in the city at the time of the first serosurvey.

Although the serosurvey was carefully designed and had adequate sample sizes, there are various possible biases which need to be considered, especially if we hope to extrapolate to the city as a whole.

1. *Possible underestimation of prevalence by igoring sensitivity of tests*. Correcting for sensitivity and specificity, the authors of [5] estimate prevalence values up to 58.3% in the slums and up to 17.1% in nonslum areas (Table 1 in the supplementary material of [5]).
2. *Possible overestimation of prevalence by choice of slums*. The largest slums in each ward were selected – this could have introduced a bias. There are reasons why, for example, a large slum might see earlier or more rapid spread of infection.
3. *Possible underestimation of prevalence by choice of wards*. The authors of [5] note that wards were selected for surveying based on the consideration that “we had NGO partners operating in the ward’s slums and the Municipal Corporation had health officers that could accompany phlebotomists as they surveyed non-slums”. By the time of the survey, the surveyed wards had generated approximately 13% fewer cases than we would expect based on even spread across the city, using the estimation process described in this technical document [15]. This suggests that these wards may not have been hit as hard as the city average by the time of the survey.
4. *Possible bias with unclear direction associated with non-consent in nonslum areas*. The authors of [5] note that in the nonslum areas, “There were difficulties in obtaining consent from some resident associations”. This could have introduced a bias if, for example, housing societies with suspected cases were reluctant to allow the surveying.

**Table 1:** Experiment 1. The values are based on 100,000 simulations of which 45,724 satisfied the inclusion criteria as described in the text. In this simulation there is no assumed drop in registration, and no drops in slum or nonslum eIFR after the first wave.

| **Experiment 1** | mean | median | 95% CI | range |
| --- | --- | --- | --- | --- |
| excess deaths | 22849 | 22881 | (21896,23728) | (21436,24083) |
| city-wide infection rate (%) | 56 | 56 | (49,63) | (43,72) |
| slum infection rate (%) | 69 | 68 | (58,81) | (54,95) |
| nonslum infection rate (%) | 43 | 42 | (32,56) | (25,73) |
| city-wide eIFR (%) | 0.32 | 0.32 | (0.28,0.36) | (0.25,0.42) |
| slum eIFR (%) | 0.28 | 0.28 | (0.24,0.33) | (0.19,0.37) |
| nonslum eIFR (%) | 0.39 | 0.38 | (0.26,0.55) | (0.19,0.78) |
| ratio of nonslum to slum eIFR (%) | 1.40 | 1.32 | (0.84,2.23) | (0.56,3.96) |
| city-wide naive IFR (%) | 0.16 | 0.16 | (0.14,0.18) | (0.12,0.20) |
| slum naive IFR (%) | 0.08 | 0.08 | (0.07,0.10) | (0.06,0.11) |
| nonslum naive IFR (%) | 0.29 | 0.29 | (0.23,0.37) | (0.18,0.43) |
| city-wide undercount factor | 2.0 | 2.0 | (1.9,2.1) | (1.9,2.2) |
| slum undercount factor | 3.4 | 3.4 | (2.8,4.3) | (2.3,4.9) |
| nonslum undercount factor | 1.3 | 1.3 | (1.1,1.7) | (1.0,1.9) |

Given the possible biases, we can check how the prevalence estimates from the first survey tally with other available data. Three other surveys give results approximately consistent with the first serosurvey.

1. *A survey in Dharavi in May-June, 2020*. According to this report in The Print [16], 36% seroprevalence was measured in Dharavi (a large slum in Mumbai) some time between May 11 and June 4. Total cases from the city’s slums, estimated following the procedure in [15], approximately doubled between June 4 and July 8. If cases tracked prevalence during this period, then 54% is not a surprisingly high estimate for slum prevalence by July 8 – if anything, it is on the low side. However, few details of the Dharavi survey are available: the data was never officially released. Dharavi may have been affected earlier than other slums. And it is not clear if Dharavi as a whole was represented, or just one containment zone within Dharavi.
2. *A survey in five slums in October 2020*. Around 75% seroprevalence was found in a serosurvey in five slums in October 2020 according to this report in India Today [17]. No technical detail is available about the sampling or the antibody test used; but in the light of these results, 54% does not seem unreasonably high for slum prevalence in early July.
3. *The second serosurvey in August, 2020* reported 45% seroprevalence in slums and 17.5% seroprevalence in nonslum areas of the same three wards as the first survey. The drop in slum seroprevalence (54% to 45%),and marginal rise in nonslum seroprevalence (16% to 17.5%), may appear at face value to indicate possible overestimation in the first survey. However, analysis shows that the numbers from the two surveys are consistent provided we take into account waning sensitivity to prior infection of the test used in both surveys. Details are given in Appendix B.

In summary, the data suggests that prevalence in slums and nonslums could have been under- or overestimated during the first serosurvey; but there is no strong evidence of bias in one particular direction. The lowest (resp., highest) values amongst *all* the 95% CI’s on prevalence values given by the authors are 52.7% (resp., 59.9%) for the slums; and 14.8% (resp., 18.4%) in nonslum areas. In later simulations we use a wider range of 50% to 62% for slum prevalence, and of 13% to 20% for nonslum prevalence, by the time of the first serosurvey.

## 5. Estimating COVID-19 infections by the end of 2020

We divide the year 2020 into two periods: *Period 1* from the beginning of the epidemic to the first serosurvey mid-point (July 8, 2020); and *Period 2* from July 8, 2020 to Dec 31, 2020. When referring to recorded COVID-19 fatalities the periods are shifted forwards by the fatality delay. For example, “Period 2 fatalities” refer to fatalities associated with added prevalence in Period 2.

We can estimate total infections by the end of 2020 by taking estimates of infections by the end of Period 1 and adding on estimates of new infections during Period 2. Since we know that detection of infections can vary substantially over time, the approach used to estimate Period 2 infections relies on fatality rather than case data. The main assumption is that the naïve IFRs in the slums and nonslum areas did not change a great deal after Period 2 began. There are reasons why these might have changed in either direction, and so in later simulations, we allow some variation in these rates.

The naïve IFRs during Period 1 are calculated by the authors of [5] (using fatality data not given in the paper) as 0.076% in the slums and 0.263% in nonslum areas. These values, if applied to the city as a whole would give a naïve IFR estimate of 0.12% at the time of the first serosurvey. They imply 2775 slum fatalities and 2573 nonslum fatalities during Period 1, assuming 54% slum prevalence and 16% nonslum prevalence and using demographic data given in Appendix A. These estimates imply that 52% of recorded fatalities during Period 1 were from the slums.

The total of 5348 Period 1 fatalities inferred from [5] is close to the 5332 COVID-19 fatalities reported in the city by July 13, 2020: thus the estimates are consistent with an assumed fatality delay of 5 days. As recorded fatalities were rising fast at the time – the total rose by 37% during July – there may be some right-censoring in these naïve IFR estimates.

In order to estimate how many new infections occurred in slums and nonslum areas during Period 2, we need first to estimate recorded fatalities from the two strata during Period 2. Before doing this systematically, let’s see an example calculation.

Suppose we fix a fatality delay – say 7 days. This gives 5698 recorded COVID-19 fatalities in Period 2 (namely, between July 15, 2020 and Jan 7, 2021). Suppose 5% of these Period 2 fatalities (285) were from the slums with the remaining 5413 from nonslum areas. Using the IFR values of 0.076% and 0.263% in [5], we would then infer 285*/*0.00076 = 3.75 × 10^5^ new slum recoveries in Period 2, and 5413*/*0.00263 = 2.06 × 10^6^ new nonslum recoveries in this period. This would amount to an additional 5.5% of slum dwellers infected, and an additional 34% of nonslum dwellers infected between July 8 and the end of year, giving end-of-year estimates of infection rate of 60% in slums, 50% in nonslum areas, and 55% city-wide.

In this calculation, the assumption that only 5% of Period 2 fatalities were from the slums is crucial. If, instead, we assumed 20% of Period 2 fatalities were from the slums, we’d get an infection rate of 61% overall (slums: 76%, nonslums: 44%) by the end of 2020. If 35% of Period 2 fatalities were from the slums, we get an infection rate of 67% (slums: 93%, nonslums: 37%) by the end of 2020.

Note, however, that there are a lot of values fixed in these estimates, including the initial estimates of slum and nonslum prevalence, the fatality delay, and the naïve IFR values. We clearly need a more systematic way of exploring such estimates, where quantities are allowed to vary over plausible ranges.

## 6. Monte-Carlo experiments

We can explore estimates of infection rates and IFR more systematically with the help of some Monte Carlo experiments. The idea is to put probability distributions on quantities where there is uncertainty (a full list is given below), and then sample repeatedly from these distributions to compute a distribution on various quantities of interest. Results of two such numerical experiments are presented. The code used is available on github [18].

From the point of view of excess deaths, we take Period 1 to comprise April, May, June and half of July 2020, and Period 2 to be the rest of the year. Including part of January 2021 in Period 2 would marginally reduce Period 2 excess deaths, as there were slight negative excess registrations during January 2021. To get death registrations for half of July we simply take half of the July total. These choices give mid-point expectations of 24659 deaths (Period 1) and 41947 deaths (Period 2), compared to actual registrations numbering 39159 (Period 1) and 50200 (Period 2). Thus, if we assume no drop in registration coverage, Period 1 saw 14,500 excess deaths, while Period 2 saw 8,253.

We allow the slum and nonslum values of eIFR to change between Period 1 and Period 2: during the first wave, the city’s health infrastructure was severely overstretched as described in this report in Article-14 [19], likely leading to preventable COVID-19 deaths, and also perhaps non-COVID deaths. Let eIFR_*s,i*_ and eIFR_*n,i*_ (*i* = 1, 2) refer to slum and nonslum eIFR during Period *i*. Excess deaths in Period *i* are then equal to

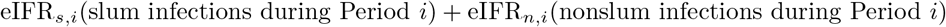

Given slum and nonslum infections during each period, excess deaths during each period, and the ratios eIFR_*s*,2_*/*eIFR_*s*,1_ and eIFR_*n*,2_*/*eIFR_*n*,1_, we can calculate eIFR_*s,i*_ and eIFR_*n,i*_ (*i* = 1, 2), and hence the excess-deaths based IFR in the city as a whole during each period and the whole of 2020.

All probability distributions used are uniform. The implications of this choice are discussed later. We use the following ranges for various parameters:

1. Variation in baseline (i.e., expected) registrations during each period: 2% in either direction. This is to allow for natural fluctuations in mortality, but without any assumption on the direction of such fluctuations.
2. (Experiment 2 only). Registration coverage drop during 2020: 0% to 5%. Even a drop of only 5% might be conservative in the light of the major drop in birth registrations.
3. (Experiment 2 only). eIFR_*s*,2_*/*eIFR_*s*,1_: 0.6 to 1; eIFR_*n*,2_*/*eIFR_*n*,1_: 0.8 to 1. I.e., we allow up to a 40% reduction in slum eIFR in Period 2, and up to a 20% reduction in nonslum eIFR in Period 2.
4. Slum prevalence at the time of the first serosurvey: 50% to 62%. Note that this range is somewhat wider than the bounds in [5] to allow for possible biases as discussed in Section 4.
5. Nonslum prevalence at the time of the first serosurvey: 13% to 20%. Again, this range is somewhat wider than the bounds in [5].
6. Percentage of Period 1 recorded COVID-19 fatalities from the slums: 0.95*52% to 1.05*52%. In other words, a 5% variation is allowed around the measured 52% (see Section 5).
7. Percentage of Period 2 recorded COVID-19 fatalities from the slums: 5% to 35%. This range is discussed in Appendix C.
8. The fatality delay: 0 days to 27 days. An integer uniform distribution is used. The high upper limit is to account for possibly lengthy delays in fatality reporting. The fact that the median estimate is higher than the 5 to 6 days calculated from [5], can be read as the assumption that there is likely some right censoring in the naïve IFR estimates given there.
9. Naïve IFR values in the slums and nonslum areas during Period 1 are not independent random variables: they are calculated from slum/nonslum prevalence and the number of Period 1 recorded COVID-19 fatalities from the slums/nonslums (see Section 5). The latter, in turn also depend also on the fatality delay.
10. Naïve IFR values during Period 2: these vary between 90% and 110% of the Period 1 values. Once a Period 1 value for, say, slum naïve IFR is fixed, we then choose the Period 2 slum naïve IFR value from between 90% and 110% of the Period 1 value. This is to allow for:
  - improved fatality recording after the first serosurvey, which would push up naïve IFR. For example, there was acknowledgement of fatality undercounting and addition of over 1,700 old fatalities in June [9]; this may have heralded a period of improved death recording.
  - a less stretched health system and improvements in treatment after the first wave passed could have pushed down naïve IFR.

We do not know in which direction such changes pushed naïve IFR, and we assume the changes were fairly modest.

### Inclusion criteria

We reject numerical experiments which result in eIFR being less than naïve IFR in either slums or nonslums, and during either Period 1 or Period 2.

The results of two sets of experiments are shown in Tables 1 and 2. The only difference is that Experiment 2 allows for some drop in registration coverage during the pandemic, and reductions in eIFR during Period 2, reflecting the improving situation in the city. Experiment 2 reflects the situation that we consider more likely.

**Table 2:** Experiment 2. The values are based on 100,000 simulations, of which 82,286 satisfied the inclusion criteria as described in the text. In this simulation upto a 5% drop in registration coverage was allowed (median drop: 2.7%). During Period 2, slum eIFR was allowed to drop by upto 40% (median drop: 22%), and nonslum eIFR was allowed to drop by upto 20% (median drop: 10%).

| **Experiment 2** | mean | median | 95% CI | range |
| --- | --- | --- | --- | --- |
| excess deaths | 25208 | 25232 | (22866,27503) | (21468,28733) |
| city-wide infection rate (%) | 59 | 59 | (51,68) | (43,83) |
| slum infection rate (%) | 74 | 73 | (60,90) | (54,113) |
| nonslum infection rate (%) | 42 | 42 | (31,56) | (25,73) |
| city-wide eIFR (%) | 0.34 | 0.33 | (0.28,0.40) | (0.24,0.49) |
| slum eIFR (%) | 0.26 | 0.26 | (0.19,0.33) | (0.10,0.41) |
| nonslum eIFR (%) | 0.49 | 0.47 | (0.28,0.76) | (0.19,1.31) |
| ratio of nonslum to slum eIFR (%) | 1.99 | 1.79 | (0.93,3.73) | (0.53,11.26) |
| city-wide naive IFR (%) | 0.15 | 0.15 | (0.13,0.17) | (0.10,0.20) |
| slum naive IFR (%) | 0.08 | 0.08 | (0.07,0.10) | (0.06,0.11) |
| nonslum naive IFR (%) | 0.28 | 0.28 | (0.22,0.36) | (0.18,0.43) |
| city-wide undercount factor | 2.2 | 2.2 | (2.0,2.5) | (1.9,2.6) |
| slum undercount factor | 3.2 | 3.2 | (2.2,4.3) | (1.2,5.4) |
| nonslum undercount factor | 1.7 | 1.7 | (1.1,2.3) | (1.0,3.4) |

Experiments 1 and 2 (Tables 1 and 2) give median city-wide infection rates by the end of 2020 of 56% and 59% respectively. Assuming some drop in registration pushes up estimates of excess mortality, particularly in nonslum areas, tending to increase nonslum eIFR.

Given the higher prevalence in the slums by the end of 2020, the median estimates from both simulations imply that in terms of excess deaths per 1000 population excess mortality in the slums and nonslum areas were comparable.

Both experiments imply a much higher ratio of excess deaths to recorded COVID-19 deaths in the slums (median slum undercount factors: 3.4, 3.2) than in nonslum areas (median nonslum undercount factors: 1.3, 1.7). The simulations confirm the intuition that the higher undercount factor during Period 1 implies that more of the city’s “uncounted” excess deaths came from the slums.

Focussing on Experiment 2, although nonslum eIFR is 79% greater than slum eIFR (median value), there is wide variance in this estimate, and we cannot say with great confidence that nonslum eIFR is definitely greater.

Histograms of the frequency of different estimates of infection rate and excess-deaths based IFR from Experiment 2 are shown in Figure 2.

**Figure 2:**
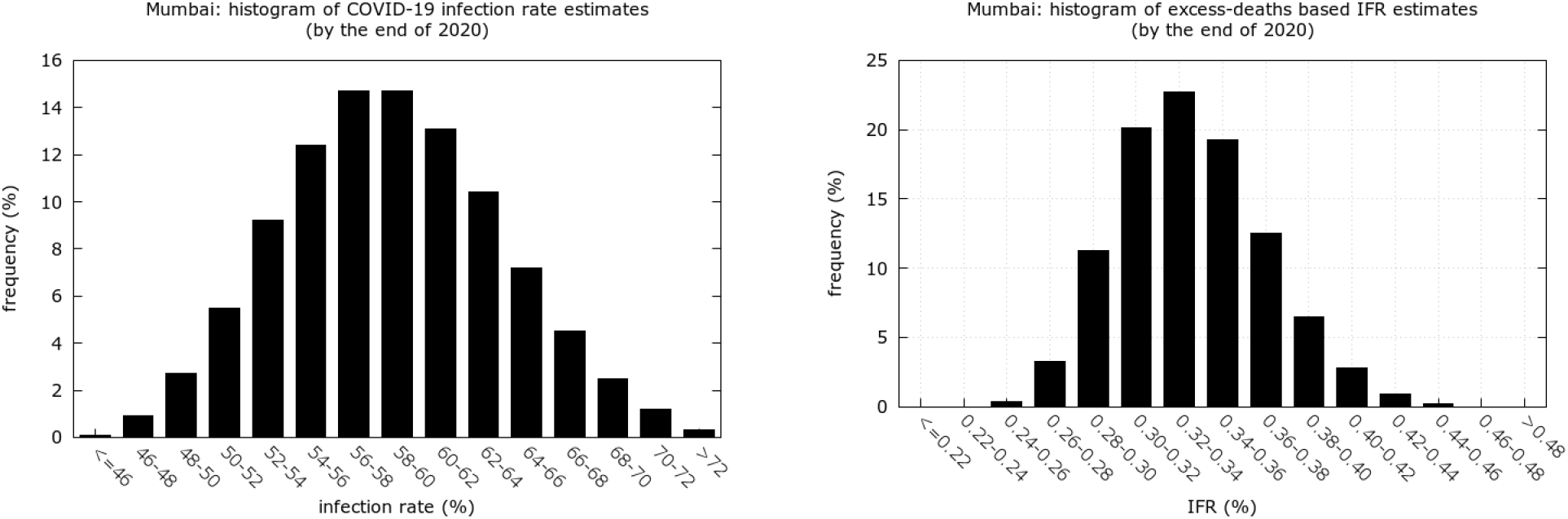
Histograms of the relative frequency of infection rate (*left*) and IFR (*right*) estimates from 100,000 experiments as described in the text.

Given the flat priors, we should interpret values with low probability as requiring a conjunction of circumstances. For example, to obtain an end-of-year infection rate below 48%, the first serosurvey must have considerably overestimated prevalence; but, in addition, there must have been relatively few subsequent infections for example if improved fatality reporting during Period 2 led to a rise in naïve IFR values.

Overall, claims that the city’s infection rate at the end of 2020 probably lies in the interval (51%, 68%), and that the city’s excess-deaths based IFR during 2020 probably lies in the interval (0.28%, 0.40%), can be regarded as statements about a balance of probabilities.

## 7. Comparison with estimates from meta-analyses

How do the IFR estimates above compare with predictions using published age-stratified data? We find that the excess-deaths based estimates lie within the range predicted using such data.

Using Mumbai’s 2011 age pyramid (see Appendix A), meta-analyses in O’Driscoll *et al* [3] and Levin *et al* [4] give estimated IFR values for Mumbai of 0.21% and around 0.37% respectively under the assumption of even spread of disease across age groups and genders.

An up-to-date age pyramid for Mumbai is not available, but moving from a 2011 age pyramid to an estimated 2021 age pyramid for Maharashtra [20], increases IFR expectations by about 35%. On the other hand, Mumbai is a city of migrants; and continuing migration into the city likely pulls the median age of the city down. We don’t know how these effects add up, but if we assume an increase in IFR of anywhere between 0% and 35% over 2011 values, we would expect values between 0.21% and 0.50% based on these meta-analyses.

### How does gender affect the estimates using [3]?

Using either gender-specific values or general values in [3] gives the same predicted IFR of 0.21% (assuming equal spread across genders). This is coincidental: the greater proportion of men in Mumbai, tending to increase predicted IFR, is offset by the lower proportion of men in the highest age groups most liable to severe infection (see Table A.3).

### Using [4] to predict IFR

The value of 0.37% computed from data in [4] uses mid-point values of the age intervals in the metaregression formula log_10_(IFR) = 3.27 + 0.0524(age), and an age of 84 to estimate fatalities for over 80s. If, for example, we replaced 84 with 82.5, this decreases the predicted IFR to 0.35%.

### Uneven spread by gender and age

The IFR estimates obtained from [3] and [4] assume equal spread across age groups and genders. But the data indicates lower spread amongst the over-60s during 2020, and possibly lower spread amongst men. Both effects would tend to reduce predicted IFR. Treating gender first, both main serosurveys showed consistently higher seroprevalence in females [21, 6] which reached significance in the slums during the first survey. Regarding age, limited demographic information in [5] (Supplementary information, Table e3) indicates that over 60s formed about 16% of the nonslum population but only about 7% of the slum population in the areas surveyed. On the other hand, median estimates of slum/nonslum infection rates by the end of 2020 indicate considerably higher infection rates in the slums. In addition, both serosurveys found significantly lower prevalence amongst the over 60s in nonslum areas. We thus expect a strong decrease in the infection rate amongst the elderly, first because the majority live in nonslum areas, and secondly because even within these areas there may have been some shielding.

## 8. Conclusions

Excess-deaths based estimates of Mumbai’s COVID-19 IFR by the end of 2020 are broadly consistent with the results of meta-analyses. Excess deaths during 2020 were around double official COVID-19 deaths, or a little more if we allow for some drop in registration coverage.

We would expect lower spread amongst the elderly, and possibly amongst men, to have decreased Mumbai’s IFR during 2020. No such decrease is clearly visible in the data. Without accurate age pyramids and mortality data stratified by age and gender it is hard to be sure, but we cannot rule out at least some avoidable deaths connected with an over-stretched health system.

The analysis tells us that the ratio of excess deaths to recorded COVID-19 deaths was likely much higher in the slums than in nonslum areas. Given inequalities in access to healthcare and testing, it is little surprise that naïve IFR estimates from the slums are likely to be more biased downwards than estimates from nonslum areas.

It is important to note that the conclusion about weaker surveillance in the slums comes from the pattern of excess mortality in the city, and without monthly mortality data it would have been very hard to arrive at this conclusion. If more granular mortality data becomes available, it should be possible to confirm directly whether the slums accounted for a much higher fraction of the city’s excess deaths than of its official COVID-19 deaths.

Finally, the city’s massive wave of infections during March-April 2021 deserves separate analysis. Data available so far is incomplete, but indicates that over the first five months of 2021, the city saw around 22% more death registrations than expected, almost all of these during April. This would mean that the 2021 surge was comparable in terms of total excess mortality to the Period 2 surge in 2020. However, the 2021 estimates of excess mortality will likely rise as more data becomes available.

## Data Availability

All data referred to in the manuscript is available publicly

## Appendix A. Notes on data used

Data on cases and recorded COVID-19 fatalities are taken from Mumbai’s daily reports from the municipal corporation. These are of two kinds: a brief report posted daily as an image on twitter; and a more detailed bulletin posted daily by the municipal corporation on the web [22]. Cumulative case and fatality data at any point is taken from the brief reports. The data is archived at [23].

The reliance on news sources for some kinds of information reflects the fact that data is often obtained by news portals but not shared publicly. The use of some estimation procedures is necessitated by the fact that data such as a breakdown of cases and deaths between slum and nonslum areas is not routinely shared; if such data were to become available, some estimates could be replaced by measured values.

2011 demographic data used for predicted IFR values is given in Table A.3. Table A.4 contains data on mortality from the Vital Statistics report [10], accessed in March 2021. Monthly all-cause mortality data in Table A.5 is taken from a variety of sources listed on github [12]. Table A.6, giving 2011 data on the slum/nonslum populations in each ward, and the city as a whole, is reproduced from [15].

**Table A.3:** Mumbai’s 2011 population by age and gender from [24].

|  | all | male | female |
| --- | --- | --- | --- |
| All ages | 12442373 | 6715931 | 5726442 |
| 0-4 | 837269 | 436703 | 400566 |
| 5-9 | 909385 | 477656 | 431729 |
| 10-14 | 998448 | 529186 | 469262 |
| 15-19 | 1100379 | 613217 | 487162 |
| 20-24 | 1333546 | 761998 | 571548 |
| 25-29 | 1310338 | 728325 | 582013 |
| 30-34 | 1109979 | 609191 | 500788 |
| 35-39 | 1015414 | 546897 | 468517 |
| 40-44 | 866960 | 466417 | 400543 |
| 45-49 | 753234 | 400590 | 352644 |
| 50-54 | 611020 | 331428 | 279592 |
| 55-59 | 481470 | 256815 | 224655 |
| 60-64 | 390770 | 200440 | 190330 |
| 65-69 | 260538 | 131235 | 129303 |
| 70-74 | 183134 | 90112 | 93022 |
| 75-79 | 110548 | 53075 | 57473 |
| 80+ | 121329 | 55159 | 66170 |

**Table A.4:** Mumbai’s estimated mid-year populations, births, all cause mortality, and infant mortality during 2015–2020, as reported in [10].

| year | 2015 | 2016 | 2017 | 2018 | 2019 | 2020 |
| --- | --- | --- | --- | --- | --- | --- |
| estimated mid-year population | 1,26,43,252 | 1,26,89,644 | 1,27,36,036 | 1,27,82,429 | 1,28,28,821 | 1,28,75,213 |
| births | 1,74,902 | 1,52,952 | 1,55,386 | 1,51,187 | 1,48,898 | 1,20,188 |
| deaths | 94,706 | 86,642 | 89,037 | 88,852 | 91,223 | 1,11,942 |
| infant deaths | 4575 | 3998 | 4079 | 3723 | 3430 | 2649 |

**Table A.5:** Mumbai’s monthly death registrations, as reported in various new sources. Sources are given on github [12]. Numbers in brackets reflect missing data, estimated from yearly totals.

|  | 2017 | 2018 | 2019 | 2017-19 average | 2020 | 2021 |
| --- | --- | --- | --- | --- | --- | --- |
| January | 8004 | 8306 | 8324 | 8211.3 | 8065 | 7732 |
| February | 7158 | 7305 | 7797 | 7420 | 7447 | 7131 |
| March | 7810 | 7436 | 7155 | 7467 | 7071 | 8302 |
| April | 6234 | 6719 | 6752 | 6568 | 7648 | 14484 |
| May | 6960 | 7407 | 7335 | 7234 | 14663 | 7514 |
| June | (7553) | 6874 | 6732 | 7053 | 11874 |  |
| July | (7553) | 7336 | 7931 | 7606.7 | 9948 |  |
| August | (7553) | 7372 | 8164 | 7696.3 | 9668 |  |
| September | (7553) | 7231 | 7953 | 7579 | 9966 |  |
| October | (7553) | (7622) | (7693) | 7622.8 | 9690 |  |
| November | (7553) | (7622) | (7693) | 7622.8 | 7614 |  |
| December | (7553) | (7622) | (7693) | 7622.8 | 8288 |  |

**Table A.6:** Total population in Mumbai’s wards at the 2011 census, broken down into those residing in slums and those not residing in slums.

| ward | total | slums | nonslums | % in slums |
| --- | --- | --- | --- | --- |
| A | 185014 | 22282 | 162732 | 12% |
| B | 127290 | 12711 | 114579 | 10% |
| C | 166161 | 16571 | 149590 | 10% |
| D | 346866 | 34699 | 312167 | 10% |
| E | 393286 | 124194 | 269092 | 32% |
| G/S | 377749 | 124306 | 253443 | 33% |
| H/W | 307581 | 82552 | 225029 | 27% |
| K/W | 748688 | 215678 | 533010 | 29% |
| R/C | 562162 | 172849 | 389313 | 31% |
| T | 341463 | 85560 | 255903 | 25% |
| F/S | 360972 | 180128 | 180844 | 50% |
| F/N | 529034 | 238128 | 290906 | 45% |
| G/N | 599039 | 361674 | 237365 | 60% |
| P/S | 463507 | 230829 | 232678 | 50% |
| R/S | 691229 | 414395 | 276834 | 60% |
| M/W | 411893 | 164992 | 246901 | 40% |
| N | 622853 | 249229 | 373624 | 40% |
| S | 743783 | 408442 | 335341 | 55% |
| R/N | 431368 | 281151 | 150217 | 65% |
| K/E | 823885 | 572818 | 251067 | 70% |
| H/E | 557239 | 388923 | 168316 | 70% |
| P/N | 941366 | 708247 | 233119 | 75% |
| L | 902225 | 758108 | 144117 | 84% |
| M/E | 807720 | 685994 | 121726 | 85% |
| total | 12442373 | 6534460 | 5907913 | 53% |

## Appendix B. Mumbai’s second serosurvey

Mumbai’s second serosurvey (approximate mid-point: August 23, 2020) was carried out in the same three wards as the first, and reported seroprevalence values of 45% in the slums and 17.5% in nonslum areas [6]. This gives around 32% seroprevalence citywide. These values appear to be adjusted for population characteristics, but not for sensitivity or specificity of the tests.

The apparent drop in seroprevalence between first and second city serosurveys is consistent with waning sensitivity of the assay used (the “Abbott Diagnostics Architect N-protein based test”) to prior infection as measured in Muecksh *et al* [7]. Calculations based on the two serosurveys, using techniques and values for the speed and frequency of seroreversion from [25] were carried out [26], and gave estimates of 28% prevalence in nonslum areas and 78% prevalence in the slums by the time of the second survey.

These estimates are somewhat higher than values predicted by tracking fatalities as in this paper, suggesting that the waning sensitivity of the tests may not have been as rapid in Mumbai as estimated from Brazilian data. Nevertheless, we can broadly conclude that the measured seroprevalence values in the first and second surveys are not inconsistent with each other.

## Appendix C. The fraction of Period 2 recorded fatalities coming from Mumbai’s slums

Using the procedure in [15], we find that around 16% of Mumbai’s COVID-19 cases between July 8 and the end of 2020 came from the slums. On the other hand, by the time of the first serosurvey, detection in the slums was much lower than in the nonslum areas, with an estimated 0.8% of slum infections identified in testing as against about 5.9% in nonslum areas [27]. If detection in the nonslum areas remained more than 7 times higher than in the slums after the first serosurvey then we can calculate, using naïve IFR estimates in slum and nonslum areas in [5], that about 29% of recorded fatalities after the first serosurvey would have occurred in the slums.

However, it is quite likely that detection of infections in the slums was particularly poor during the massive slum surge in April and May and improved (proportionally) more significantly than detection in the nonslum areas during Period 2. If, say, slum detection tripled during Period 2, while nonslum detection only doubled, then around 21% of Period 2 recorded fatalities would have occurred in the slums.

This appears a plausible scenario, but given the uncertainties, we allow a wide range: between 5% to 35% of Period 2 COVID-19 deaths occurred in the slums. The lower figure corresponds to detection in the slums achieving parity with detection in nonslum areas during Period 2. The upper figure corresponds to detection in nonslum areas being almost 10 times higher than in the slums during Period 2.

